# Navigating Care in Crisis: A Qualitative Study of Healthcare Access Among Ethnically Diverse COVID-19 Patients in The Netherlands

**DOI:** 10.64898/2026.07.10.26357237

**Authors:** Nikita Hensen, Gioia Nydia Muru, Maria Prins, Karien Stronks

## Abstract

Ethnic minority and migrant populations experienced disproportionately severe COVID-19 outcomes across Europe, yet the mechanisms underlying these disparities, particularly inequities in healthcare access, remain insufficiently understood at the patient level. This qualitative study examines healthcare-seeking behaviours and access to care among ethnically diverse patients hospitalised with COVID-19 in Amsterdam between 2020 and 2022, and the contextual factors shaping their pathways to care.

Twenty adults of Turkish, Moroccan, Surinamese, Ghanaian, and Dutch ethnic backgrounds, all hospitalised with COVID-19, were interviewed using a semi-structured retrospective approach to reconstruct individual care pathways from symptom onset to hospitalisation. Data were analysed thematically, guided by the Candidacy Framework and the Health Belief Model.

Pandemic-induced structural disruptions, including healthcare system strain, capacity shortages, absent care protocols, and fragmented referral pathways, constituted the primary barriers to care across all ethnic groups. Participants with longer hospital stays tended to be older, less educated, and with more comorbidities, yet reported fewer barriers once hospitalised, as disease severity triggered prioritisation. Those with shorter stays or emergency department visits without admission encountered greater difficulties, including repeated discharge despite worsening symptoms. Language barriers and prior negative experiences with healthcare services compounded access challenges for some participants with migrant backgrounds, though pandemic phase and disease severity were the dominant determinants across the sample.

Inequities in access to care were driven primarily by pandemic-induced structural factors rather than ethnic background. Pre-existing vulnerabilities among migrant groups, including reduced institutional trust and language barriers, intensified these structural barriers for some. These findings are directly relevant for equity-sensitive pandemic preparedness: crisis response frameworks must explicitly address structural accessibility alongside targeted support for groups facing compounding disadvantage.

## Introduction

The COVID-19 pandemic has brought renewed attention to the existing health disparities between various population groups. A systematic review investigating coronavirus disease 2019 (COVID-19) outcomes in the European Region of the World Health Organization (WHO) showed that incidence, risk of severe disease, hospitalisation rates and mortality were all higher in migrant groups and in ethnic minorities in comparison with levels in majority populations (1). Data from several countries indicate that such adverse outcomes likely resulted from complex interplays between socioeconomic disadvantage (which increased exposure to SARS-CoV-2) and a higher burden of pre-existing morbidities that predisposed individuals with migrant backgrounds to more severe illness (2–5). Evidence suggests that differences in healthcare-seeking behaviours, defined as the actions individuals take to resolve their health problems, contribute to health disparities between population groups (6, 7). Such behaviours not only affect health outcomes but also shape the pathways individuals take through the healthcare system, directly influencing their utilisation of health services (8). Moreover, the behaviours reflect and interact with inequalities in the accessibility of healthcare systems. For example, data from the HELIUS study by Amsterdam UMC and the Public Health Service of Amsterdam has revealed that people with backgrounds of migration from lower-income countries were less likely to get tested and vaccinated for COVID-19 than residents of ethnic Dutch origin (9, 10). This suggests that barriers in access to care, such as delays in seeking healthcare for COVID-19, may be more numerous in such minority groups than in majority populations (4, 11, 12). Research indicates that access to care may be influenced by a combination of economic, legislative, cultural, social, and health system factors, which vary across and between ethnic groups (13–15). In addition, the pandemic exacerbated the challenges posed by structural racism and social influences (16), and that could have interacted with implicit biases in healthcare providers (17). As a consequence, minority ethnic patients could have received less timely or adequate medical care and treatment during the pandemic than other patients. Although such findings suggest disparities between ethnic groups in access to care, there is hardly any empirical evidence on whether differences in health system accessibility might have underlain the adverse outcomes of COVID-19 as reported in specific population groups. From the perspective of pandemic preparedness, exploring access to healthcare in such populations, and the factors influencing that access, is crucial for addressing healthcare disparities in future pandemics (18). The aim of this paper is to study accessibility of care and related healthcare-seeking behaviours among people from different ethnic groups who were hospitalised with COVID-19, as well as the factors that shape such accessibility and behaviours. Central to this paper are the following research questions:

1. What healthcare-seeking behaviours, and related barriers to accessible care, can be observed in ethnically diverse patients admitted to hospital with COVID-19?
2. What contextual factors influence the healthcare-seeking behaviours and the access to COVID-19–related care in patients from different ethnic backgrounds?

## Materials and methods

### Design

This study adopted an individual semi-structured interview approach to comprehensively investigate healthcare-seeking behaviours and access to care in individuals admitted to hospital with COVID-19. The interviews were retrospective in nature, reconstructing the disease course leading to hospitalisation in order to ensure a thorough examination of the participants’ experiences (see S1 Supporting Information). The study received ethical approval from the Medical Ethics Review Board (METC) of Amsterdam UMC, which also determined that the study was outside the scope of the Medical Research Involving Human Subjects Act (WMO) of the Netherlands.

### Population and recruitment

The study sample consisted of adult Amsterdam residents of Turkish, Moroccan, Surinamese, Ghanaian and Dutch ethnic origin who had been hospitalised with COVID-19 between 2020 and 2022. Those groups were selected as representing Amsterdam’s largest ethnic populations and because of the reported higher hospitalisation rates among individuals with non-European migrant backgrounds (19). Participants were recruited through the Healthy Life in an Urban Setting (HELIUS) study, a prospective cohort study investigating health disparities in Amsterdam’s multiethnic population. Eligible individuals who reported COVID-19 hospitalisations in the HELIUS COVID-19 sub-study (5) and consented to further research were approached by phone and invited to participate in interviews between April and June 2023. Of 44 eligible participants, we were able to reach 37, and 20 of these agreed to participate, with the main reason for declining being lack of time (*n* = 11). Interviews were scheduled based on participant availability, and transport vouchers were provided to cover travel expenses. Purposive sampling ensured diversity across the participant groups.

### Public involvement statement

Participants were not directly involved in the design, conduct, or dissemination planning of this sub-study. However, the research was embedded within the HELIUS study, which has prioritised community engagement since its inception. This engagement has informed the development of research questions, culturally sensitive study conduct, and dissemination strategies. The current study was shaped by this broader participatory framework.

### Context

In response to the COVID-19 pandemic, the Netherlands implemented various mitigation measures that influenced daily life. The evolving understanding of the virus over time, combined with the introduction of vaccinations, shaped the healthcare landscape. Patients seeking hospitalisation at different times might have experienced different contexts, from initial uncertainties and fears to later stages marked by increased knowledge and reduced uncertainty (20).

### Data collection

Data collection took place from May to June 2023, either in a private room at Amsterdam UMC, online via Teams, or at the participant’s home, depending on participant preference. During that period, none of the former Dutch COVID-19 measures, including those regarding testing and isolation, were still in place (20). All interviews were conducted by NH, a female junior researcher with an MSc in Health Sciences, affiliated with the Department of Public and Occupational Health at the time of the study. NH had prior experience with semi-structured interviewing and had received formal training in qualitative interviewing techniques. Assistance by interpreters was provided if participants preferred communication in their native language (*n* = 3). No prior relationship existed between the researcher and participants, and participants were not informed of the researcher’s personal goals or motivations. All interviews were audiotaped and transcribed. Interview duration ranged from 45 to 120 minutes. Verbal and written informed consent were obtained from participants prior to the interviews, with anonymity in the presentation and publication of data assured. The final interviews did not yield additional themes, indicating that data saturation had been reached.

### Data analysis

Thematic analysis was applied, and an initial coding system was developed based on the theoretical framework, published literature and preliminary insights (see S2 Table). A combination of deductive and inductive coding strategies was used to systematically analyse the data. The coding process was led by the first author, using MaxQDA (V. 2018, VERBI, Berlin), with iterative adjustments made to the coding scheme as new themes emerged. NH and KS held regular discussions to refine emerging themes, patterns and results. To further enhance the credibility of the findings, GM conducted a coding check across a subset of eight interviews with regard to the most challenging themes, including identification of candidacy, permeability of healthcare services, and patients’ appearances at services. Additionally, GM performed a textual quality review for all themes.

### Conceptual models

The concept of candidacy (21) served as our primary framework for exploring participants’ healthcare-seeking behaviours and experiences before and during COVID-19 hospitalisation. Candidacy is understood as a dynamic concept that is continuously shaped and negotiated by factors such as personal circumstances, socioeconomic conditions, societal structures, and availability of resources. Liberati et al. (2022) demonstrated the utility of the candidacy construct in highlighting how service reconfigurations during the pandemic affected help-seeking behaviours and healthcare accessibility, suggesting refinements to the original model (22). In response, we updated its features to reflect the COVID-19 context, as shown in Table 1. We also incorporated elements of the Health Belief Model (HBM) (23), which provided insights into participants’ perceptions of disease severity and susceptibility and of barriers to care. By integrating the two models, we sought to gain a nuanced understanding of the interplay between participants’ health behaviours and the interactions within the affected healthcare system.

**Table 1.**
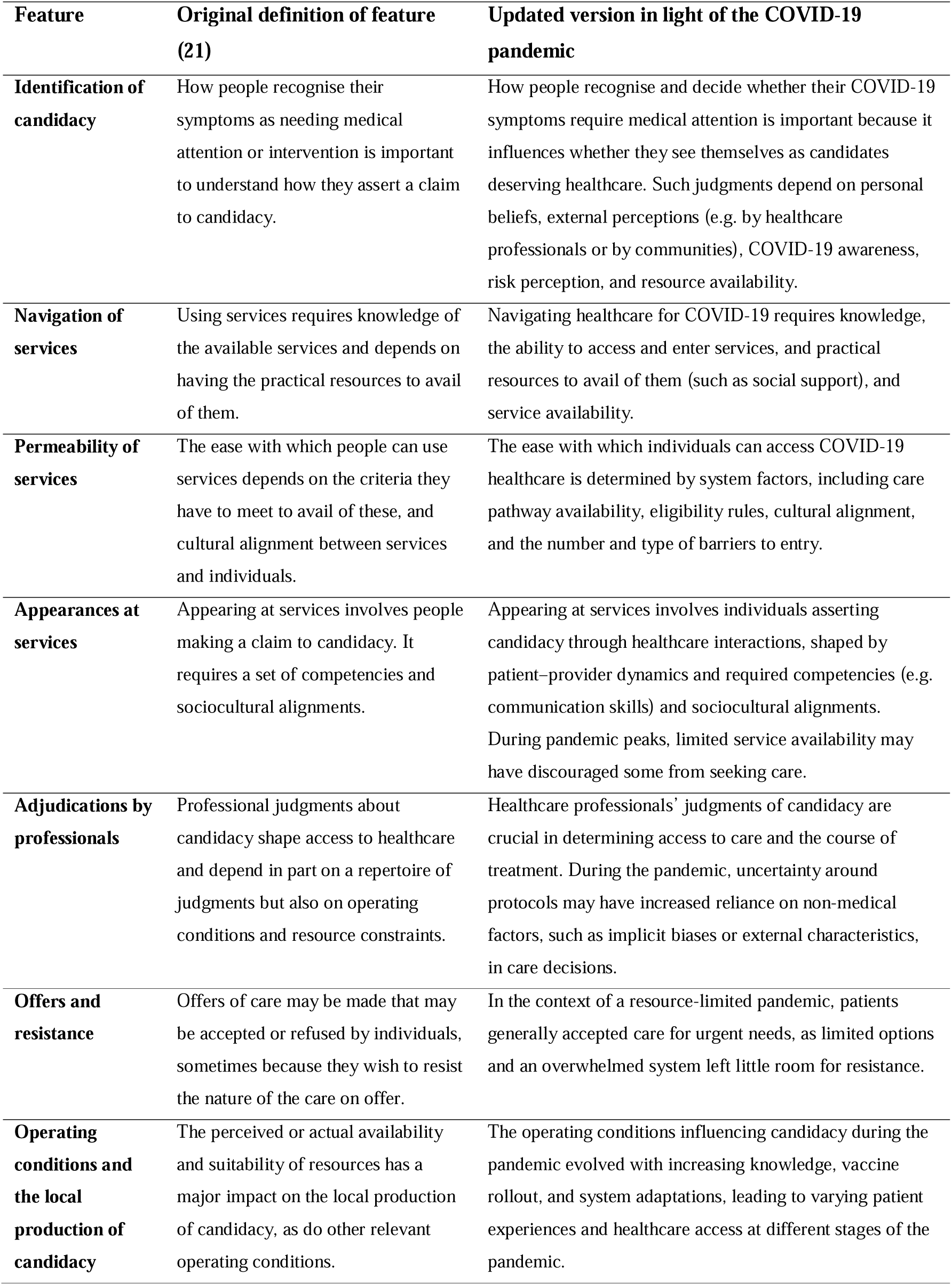
Understanding candidacy for COVID-19 healthcare.

## Results

Interviews were conducted with a total of 20 participants of diverse ethnic origin, including nine Surinamese, five Turkish, two Moroccan, three ethnic Dutch, and one of other ethnic background. Two thirds were male (*n = 13*); most participants were aged between 60 and 69 years (*n = 13*); and most had lower to intermediate education levels (*n = 13*). Common comorbidities included cardiovascular diseases (*n = 8*), with several respondents managing multiple conditions such as diabetes or asthma (see Table 2 for participant characteristics). This study focused on COVID-19 patients hospitalised at some point during the pandemic, examining their healthcare trajectories before and during hospitalisation. We grouped participants based on the type and duration of their hospital stays: (1) ICU patients with a prolonged stay (>14 days, *n = 4*); (2) longer hospital stays (5–13 days, *n = 7);* (3) shorter stays (1–4 days) and (4) emergency department visits without admission (total *n = 9*). In Table 2, groups 3 and 4 are presented as a combined category to prevent identification of participants given the small number in group 4. Participants in groups 1 and 2 tended to be older, male, and less educated, and most had been hospitalised during the early or peak phases of the pandemic. In contrast, group 3 and 4 participants were more often younger, female, more educated, and treated during later or non-peak periods. In the sections to follow, we describe how structural and contextual factors shaped healthcare-seeking behaviours and access to care across groups, using the Candidacy Framework as an analytical lens.

**Table 2.**
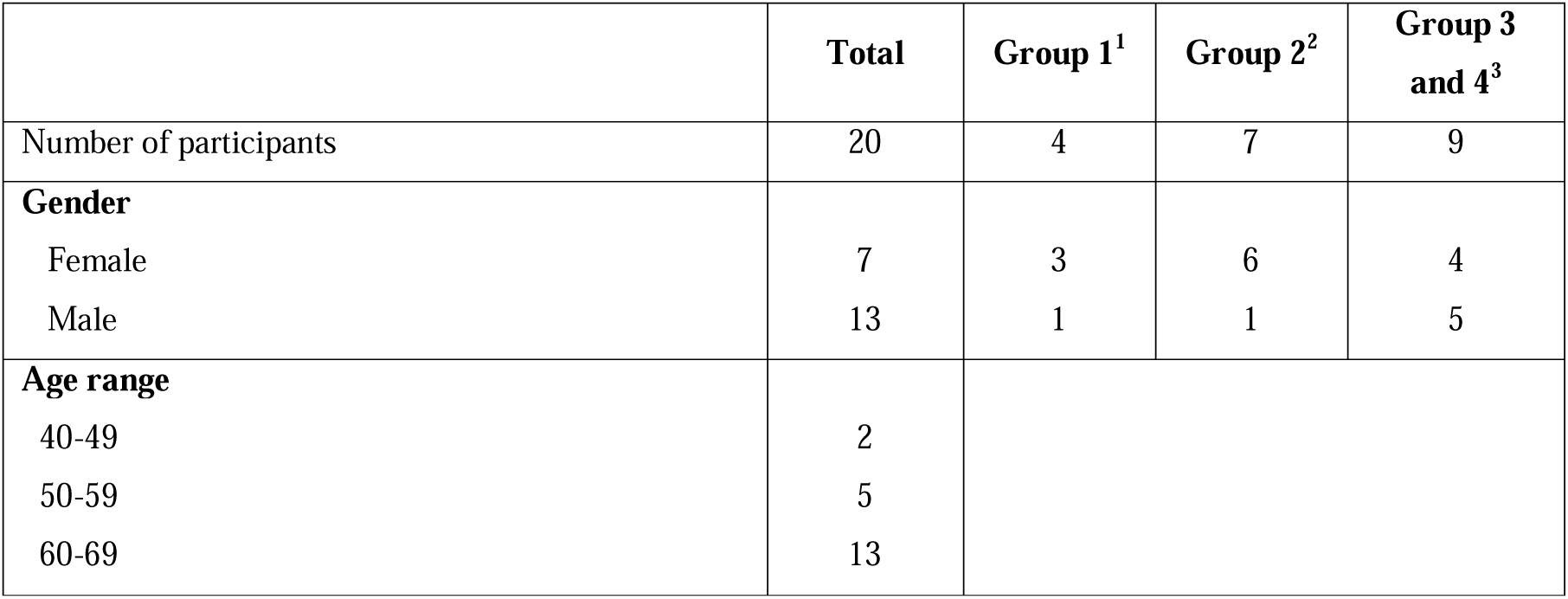
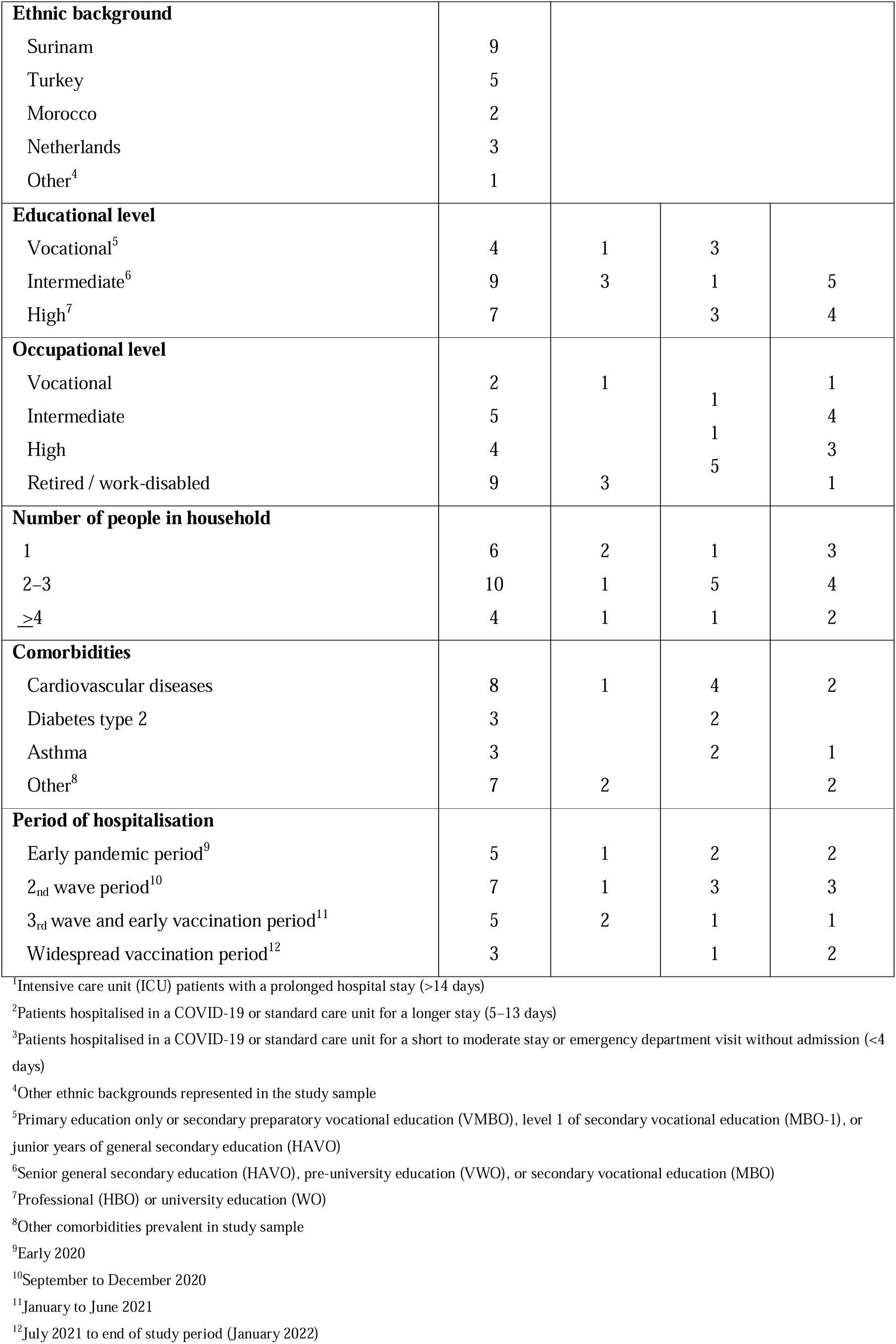
Sample characteristics.

### Identification of candidacy

Participants’ decisions on whether their symptoms required medical attention depended notably on the perceived severity of their problems and the extent to which they associated their symptoms with COVID-19. The observed patterns remained consistent across ethnic groups within each category of hospitalisation duration. Participants who contracted COVID-19 during the first waves of the pandemic often underestimated their symptoms due to the novelty of the virus. As public awareness and knowledge grew over time, participants became more likely to link their symptoms to COVID-19. Most participants reported that their initial symptoms were similar to those of common cold or flu. Persistent or worsening symptoms eventually led them to seek help from their general practitioner (GP), thus recognising their need for medical attention. However, in most cases their need for urgent care did not get fully recognised initially and they were sent home. Some also provided examples of misinformation circulating within their communities.

> In the early days of the pandemic, I received a lot of advice on how to prevent COVID. There was so much bizarre information online – some people suggested using a blood donor, while others said to stop drinking alcohol. I didn’t believe any of it. [P13: Turkish background, group 2]

Participants admitted to the ICU for prolonged stays (group 1) experienced rapid disease progression. Two of them were unaware it was COVID-19 and collapsed, falling directly into a coma. Ultimately, all participants in this group were too ill to seek help themselves and had to rely on bystanders or family members to summon an ambulance. Once hospitalised, participants reported that they were unable to communicate their care needs. Due to the critical nature of their condition, they were sedated, and hospital staff assumed full responsibility for their care. Some participants admitted to standard care wards for varying durations (groups 2 and 3) or those who applied to the emergency department but were not hospitalised (group 4) reported delays in receiving care or being sent home prematurely, often necessitating a return to hospital. Their decisions to seek medical care prior to hospitalisation were driven by deteriorating symptoms, encouragement or assistance from others, and their COVID-19 risk perception. Those who did not perceive themselves at high risk, particularly younger participants and those without comorbidities, more often reported not seeking medical care until their condition became critical.

> Being young and living a healthy lifestyle, I didn’t consider myself at high risk. At first I thought, like most people, ‘It’s probably nothing serious.’ But that’s not true – COVID-19 can have very serious consequences for anyone, no matter what their background or lifestyle. [P12: Moroccan background, group 2]

Participants in group 4 reported feeling a lack of control over their situation. Although they recognised the need for medical assistance due to severe symptoms, such as difficulty breathing, they were sent home without care, leaving them uncertain about what to do next.

> I was sent home by my GP despite my symptoms. It felt like I was dying – like I was sinking into quicksand, wanting to reach out but just sinking deeper. Thankfully, I made it through. [P11: Surinamese background, group 4]

### Navigation of services

Participants sought healthcare at testing facilities, GP offices, hospital emergency departments, or a combination of these, often relying on government resources to guide their decisions. Across all ethnic groups, disease severity and the stage of the pandemic were the main factors influencing how participants accessed care and the resources they needed. Participants who became ill during the early waves of the pandemic frequently reported feeling uncertain about the appropriate actions to take. Most participants initially sought help from their GP and later accessed hospital care through ambulance transport or GP referrals. Many relied on social connections for support and translation when navigating for care. However, some participants related how lockdown measures limited access to such networks, thus exacerbating feelings of isolation and uncertainty. This applied especially to those with language barriers, who highlighted the need for clear COVID-19 care routes.

> It’s very difficult for me to understand Dutch. I normally acquire information from the people around me, but due to the government constraints, I couldn’t. In a crisis like COVID-19, information should be more easily accessible and provided in multiple languages. [P18: other ethnic background, group 1]

Two participants eventually admitted to the ICU for a prolonged stay (group 1) had had to repeatedly navigate to their GP to get hospitalised. All participants in group 1 suffered severe symptoms that required emergency ambulance transport, leaving medical professionals in charge of their care pathways. Participants with longer non-ICU hospital stays (group 2) reported severe disease progression that led to their prioritisation over others with less critical conditions. Those in group 3 and 4 had had difficulty reaching their GP and many had been sent home despite worsening symptoms. Although this challenge was not specific to migrant backgrounds, those who struggled with the Dutch language shared how such experiences further complicated their access to care, leaving them feeling hopeless and uncertain about the next steps.

> During the COVID period, we were completely unable to reach our GP. Despite being extremely ill, I couldn’t get in touch with them by phone or in person. It felt as though someone with heart attack were left to suffer without help. It was a very harsh and distressing experience. [P14: Turkish background, group 2]

Participants often required additional GP visits or ambulance calls before arriving at a hospital emergency department, where some faced long waiting times before being admitted or even sent home on grounds of capacity management. Those sent home to recover independently (group 4), often felt uncertain about where to seek further help, but some proactively pursued physiotherapy to aid their recovery. Other participants (in groups 1 to 4) reported hesitance to seek medical care due to past negative experiences, such as negligence by health professionals and feeling ignored in medical settings. Such experiences were most frequently reported by those with migrant backgrounds, affirming that this raised their threshold for seeking care. Others delayed seeking care until their condition became critical, fearing they might overburden the strained healthcare system.

> I was hesitant to contact the doctor about my symptoms because the doctors were already so overburdened with work. I thought it was all just inconvenient for others. I can solve it myself or not, or it will pass. I didn’t want to be a burden… [P4: Dutch ethnic background, group 3]

### Service permeability

Participants’ ability to access appropriate care for COVID-19 symptoms was shaped by accumulating challenges, from limited knowledge early in the pandemic to capacity constraints and resource shortages during peak waves. A key barrier for all groups was the fragmented nature of the healthcare system during the pandemic, which led to frequent referrals, disrupted care continuity, and strained patient–doctor relationships. These disruptions often caused delays in diagnosis, miscommunication, and reduced trust due to non-familiarity with healthcare professionals, independent of ethnic background.

> It was difficult with a substitute doctor who didn’t know me, and I think, had it been my own doctor, things might have gone faster! [P10: Surinamese background, group 3]

Participants admitted to an ICU (group 1) experienced fewer barriers once hospitalised, due to prioritisation of their critical condition. However, participants who initially visited their GP faced challenges convincing them of their need for hospital admission. One participant recounted multiple GP visits in which he was sent home with paracetamol until his condition critically deteriorated, eventually requiring ambulance transport and a month-long sedation in the ICU. Participants admitted to hospital, but not to the ICU (groups 2 and 3), reported delays in diagnosis due to the absence of well defined care pathways and protocols for COVID-19.

> My GP was upfront, saying, ‘I’ve never seen this before.‘ It was unsettling because it made me worry that ‘if they don’t know what’s happening, how will they help me?‘ I didn’t feel safe at all; it felt like I could collapse at any moment. The uncertainty was terrifying! [P10: Surinamese background, group 3]

Participants hospitalised for shorter periods often mentioned being discharged before full recovery, while those with longer stays reported challenges with multiple hospital transfers due to capacity constraints and prioritisation of more critical patients. In all ethnic groups, many struggled to understand their medical conditions and treatment options, citing poor communication and inconsistent information from healthcare providers. Some experienced varying levels of care across hospitals, attributed to factors such as time constraints and perceived lack of expertise during consultations. Such issues left participants, especially those facing language barriers, feeling frustrated and less autonomous in managing their care.

> Communicating with doctors is challenging for me. When I go alone, even if I try to say things, I often forget what I wanted to say. I have to think in Turkish, translate it into Dutch, and then try to respond. This constant switching between languages makes it really difficult to ask questions and express my concerns clearly. [P14: Turkish background, group 2]

Participants who had visited an emergency department but were not hospitalised (group 4) experienced the least permeable healthcare pathways. They encountered barriers similar to those of others but lacked the resources or perseverance to overcome them, repeatedly experiencing ineffective treatment and follow-up. One participant voiced frustration at being misdiagnosed early in the pandemic, despite presenting clear COVID-19 symptoms.

> No one seemed to make the connection. It was ‘still pneumonia’. No, I hadn’t been to China and, no, I hadn’t eaten bat meat. The questions were just absurd. Anyway, I’m glad it eventually passed, but I do have a lasting spot on my lung. [P11: Surinamese background, group 4]

### Appearance at services

The extent to which participants claimed candidacy in their interactions with healthcare providers depended largely on the availability of healthcare services during the pandemic. Participants explained how peaks in COVID-19 cases confronted them with multiple challenges, including hospital transfers, early discharges, and restrictive measures to prevent virus transmission. Such restrictions often hampered participants’ access to support from family and friends.

> When I was in hospital, only one person was allowed to visit due to COVID-19. There you are, lying there with a death sentence, and then you’re not even allowed to receive visitors. I found it a really terrible time. [P4: Dutch ethnic background, group 3]

Some participants explained that the lack of support from their relatives hindered their ability to advocate for care, as it made understanding medical information more difficult and heightened their distrust of the healthcare system.

Participants with prolonged stays in intensive care (group 1) explained that, after regaining consciousness and beginning their recovery, the significant physical and mental toll of their illness made it difficult to advocate for themselves. Despite such challenges, all participants in this group expressed satisfaction with the care they received, emphasising that healthcare providers had kept them well informed about their condition. Participants from groups 2 to 4 reported varying experiences with providers at both GP and hospital levels. Many participants felt supported by providers who listened attentively, particularly at the GP level, where an existing relationship of trust often facilitated better communication and understanding. In contrast, interactions in hospitals, especially during pandemic peaks, were often perceived as impersonal, leaving participants feeling unheard and disconnected.

> They [the hospital staff] could have asked me more about how I was feeling and if I was truly ready to go home. It felt like they just wanted to get rid of me. They should have taken the time to understand my thoughts and feelings better. [P7: Turkish background, group 2]

The strain on healthcare resources during the pandemic influenced discharge decisions. Some participants were sent home earlier than they felt ready, in order to accommodate more critical patients, whilst others, feeling well enough, requested early discharge to avoid adding to the burden on the already overwhelmed system.

### Adjudications by providers

Participants across all groups shared examples of how the pandemic context and resource constraints influenced healthcare professionals’ judgments, leading to what some perceived as moral injury. A few participants mentioned their impression that critical cases were prioritised over less severe ones, which may have also impacted referral and admission processes for some patients. While participants did not attribute those challenges to sociocultural factors, many believed that the healthcare system and providers were unprepared to effectively treat COVID-19. Those hospitalised later in the pandemic reported more satisfaction with the decisions of their healthcare providers, though some continued to experience the effects of strained services. Ethnicity did not appear to affect providers’ adjudications, and most minority ethnic participants did not feel marginalised. However, one participant pointed out difficulty of engaging with healthcare professionals due to language barriers.

> The other person just doesn’t want to listen. You ask a few questions, but if you don’t express yourself perfectly, they give up quickly because they don’t fully understand. And then you start feeling like an outsider – excluded, useless. It weighs heavily on you. [P14: Turkish background, group 2]

Despite these challenges, most participants expressed understanding of the immense pressure on healthcare providers during the pandemic, admiring their efforts under difficult conditions.

> It was complete chaos at the time. I was in the inpatient ward, and there were people denying that anything was wrong. Meanwhile, I could hear quarrels between patients and nurses, and some visitors weren’t even wearing masks. It created a very tense atmosphere. I can imagine how frightening it must have been for the nurses working there, too. [P4: Dutch ethnic background 3]

### Offers and resistance

Amid the pandemic context, most participants, independent of ethnic background, assented to the care offered, particularly when it addressed urgent needs. Those with critical conditions often had no choice but to accept treatments due to the severity of their symptoms, the lack of specific COVID-19 treatments, and the strain on healthcare resources. In contrast, participants with milder symptoms faced greater difficulty accessing appropriate care, often accepting the limited treatment options available, uncertain about how to navigate their healthcare in such constrained circumstances.

## Discussion

### Key findings

This study examined healthcare-seeking behaviours and access to care among ethnically diverse patients hospitalised with COVID-19 in Amsterdam between 2020 and 2022, focusing on the structural and contextual factors shaping their pathways to care. The findings reveal that pandemic-induced structural barriers, such as healthcare system reconfigurations, capacity management decisions, and the absence of established care protocols, were the primary drivers of inequities in access to care, operating across all ethnic groups. Participants hospitalised during the same pandemic phase reported strikingly similar barriers, regardless of ethnic background, and these structural conditions shaped how care was delivered, how priorities were set, and how resources were allocated. The limited permeability of healthcare services emerged as a key mechanism affecting access, particularly at the initial point of contact, where systemic unpreparedness and dynamic capacity constraints created barriers for all patient groups. These structural disruptions intersected with, rather than were superseded by, individual social determinants, particularly language barriers and reduced institutional trust, which compounded the challenges faced by some participants with migrant backgrounds.

### Interpretation of findings

The profile of participants with prolonged hospitalisations (older, less educated, with higher comorbidity burdens) is consistent with established risk factors for severe COVID-19 outcomes (24), suggesting that disease severity, rather than healthcare-seeking behaviour or ethnic background, was the primary determinant of hospitalisation duration and care trajectory. Hospitalisations in this group occurred predominantly during the early, high-pressure phases of the pandemic, characterised by high infection rates and limited system capacity (25). Although all participants recognised their symptoms and actively sought care, the strain on the healthcare system shaped the boundaries of candidacy in practice: participants whose symptoms did not meet the threshold for critical prioritisation found their care needs consistently unmet, irrespective of their healthcare-seeking efforts. This pattern suggests that pandemic-induced capacity constraints created a de facto inequity in care options, disproportionately affecting those with significant but non-critical symptoms, often younger, more educated participants who nonetheless struggled to obtain appropriate care despite actively seeking it.

In addition, social networks played a critical role in supporting healthcare-seeking across all groups (26, 27) though lockdown measures limited access to these networks for some, exacerbating feelings of isolation and uncertainty, particularly among those with language barriers. Concerns about burdening an overwhelmed system contributed to delays and care avoidance across all ethnic groups (24, 28). As the pandemic progressed, improved access to testing, vaccinations, and care protocols reduced hospitalisations and eased system burden (29, 30); however, during peak periods healthcare systems remained overwhelmed, limiting access for non-urgent patients and causing widespread delays (31, 32).

Furthermore, participants consistently reported difficulties at their first point of contact, such as frustration with brief doctor consultations and perceived uncertainty of health professionals on how to diagnose and manage COVID-19 cases, particularly during the early waves. As a consequence, many participants were sent home and experienced delays in receiving appropriate care, independent of ethnic background. The fragmented nature of the healthcare system caused disrupted referral pathways, diagnostic uncertainty, and strained patient-provider relationships, fostering fears of not being taken seriously and delaying treatment (33, 34). Participants with migrant backgrounds, particularly those facing language barriers, described additional challenges in self-advocacy, including difficulties articulating symptoms and understanding medical information, compounding the structural barriers shared by all participants (35). More broadly, the impersonal nature of pandemic-era care discouraged patients from asking questions or advocating for themselves, limiting understanding of medical decisions and reinforcing perceptions of poor communication (36). Stress and burnout among healthcare workers may have further exacerbated such challenges (37).

Participants’ perceptions of the healthcare system also shaped their care-seeking, with some expressing criticism of the Dutch pandemic response despite acknowledging the care they received (38). Prior negative experiences can foster distrust, lowering COVID-19 risk perception and compliance with public health measures (39, 40). For some participants with migrant backgrounds, this distrust was rooted in systemic inequalities and past negative encounters, potentially contributing to delayed care-seeking (41, 42). These findings point to a structural inequity embedded in the pandemic response itself: a system under strain that created new barriers for all patients while simultaneously intensifying pre-existing vulnerabilities for those with migrant backgrounds. Addressing access inequities in future crises therefore requires both system-level resilience and targeted support for groups facing compounding disadvantage.

### Limitations

Several limitations warrant consideration. Selection bias may have affected representativeness, as only COVID-19 survivors who consented to participate were included, potentially excluding those with the most severe or fatal experiences. The study was geographically confined to Amsterdam, which may limit generalisability; however, Amsterdam’s ethnic diversity and the universal nature of pandemic-related healthcare challenges suggest relevance to comparable urban settings. The study captures patient perspectives only; incorporating healthcare provider viewpoints could have added depth to understanding system-level dynamics. Recall bias is a potential concern given that interviews were conducted one to three years after hospitalisation, though the life-altering nature of COVID-19 and the consistency of reported experiences across participants suggest key details were well retained. Finally, most interviews were conducted by an ethnic Dutch researcher, with interpreter assistance in three cases. The researcher’s background relative to the diverse participant group may have introduced social desirability bias and affected depth of disclosure, particularly on topics related to experiences of discrimination or institutional mistrust. Efforts to create a safe interview environment, including emphasising confidentiality and allowing participants to choose their preferred format, may have helped to mitigate these concerns.

### Implications

These findings carry direct relevance for equity-sensitive pandemic preparedness, a policy priority across Europe and globally. Comparative analyses of European pandemic preparedness plans have highlighted a persistent absence of equity considerations for marginalised groups (43, 44); this study provides the patient-level qualitative evidence needed to address that gap. Three implications stand out. First, healthcare systems must build structural resilience that does not reproduce inequities under pressure: capacity management decisions, referral protocols, and discharge policies all carry equity consequences that must be explicitly considered in crisis planning. Second, communication strategies during health emergencies must be proactive, multilingual, and culturally accessible, not only to reach minority populations, but to maintain the institutional trust on which timely care-seeking depends. Third, the Candidacy Framework proved a valuable lens for making visible the dynamic and negotiated nature of care eligibility during a crisis, and offers a useful tool for designing and evaluating equitable crisis response strategies.

## Conclusion

Access to care among ethnically diverse patients hospitalised with COVID-19 was shaped primarily by pandemic-driven structural factors, including system overload, capacity management decisions, and fragmented care pathways , rather than by ethnic background. These structural conditions affected all patient groups, while simultaneously intensifying pre-existing vulnerabilities, particularly language barriers and reduced institutional trust, for some patients with migrant backgrounds. As pandemic preparedness frameworks are actively being developed across Europe, embedding equity as a core design principle, rather than an afterthought, is essential to prevent the reproduction of these disparities in future health emergencies.

## Data Availability

The data that support the findings of this study are available on reasonable request from the corresponding author. The data are not publicly available due to privacy and ethical restrictions.

## Acknowledgements

The HELIUS study, a collaboration between Amsterdam UMC and the Public Health Service of Amsterdam (GGD), is conducted with core support from both organisations. We sincerely thank the HELIUS participants, the management team, and the interpreters who facilitated data collection for this research. We also sincerely thank M. Dallas for his valuable editorial support in refining the language and clarity of this manuscript.

## Supporting Information

**S1.** Interview topic list

**S2 Table**. Coding framework

